# Antibodies targeting merozoites induce natural killer cell degranulation and interferon gamma secretion and are associated with immunity against malaria

**DOI:** 10.1101/2021.12.14.21267763

**Authors:** Dennis O. Odera, James Tuju, Kennedy Mwai, Irene N. Nkumama, Kristin Fürle, Timothy Chege, Rinter Kimathi, Stefan Diel, Fauzia K. Musasia, Micha Rosenkranz, Patricia Njuguna, Mainga Hamaluba, Melissa C. Kapulu, Roland Frank, Faith H. A. Osier, CHMI-SIKA Study Team

## Abstract

Natural killer cells are potent immune effectors that can be activated via antibody mediated Fc receptor engagement. Using multiparameter flow cytometry, we found that natural killer (NK) cells degranulate and release IFNγ upon stimulation with antibody-opsonized *Plasmodium falciparum* merozoites. Antibody-dependent NK activity (Ab-NK) was largely strain-transcending and enhanced the inhibition of invasion into erythrocytes. Ab-NK was associated with the successful control of parasitemia following experimental malaria challenge in African adults. In an independent cohort study in children, Ab-NK increased with age, was boosted by concurrent falciparum infections and associated with a lower risk of clinical episodes of malaria. Nine of 14 vaccine candidates tested induced Ab-NK including some less well-studied antigens - P41, P113, MSP11, RHOPH3, and *Pf*_11363200. These data highlight an important role for ab-NK in immunity against malaria and provide a new mechanism for the evaluation of vaccine candidates

**One Sentence Summary:** Antibody-dependent natural killer activation is induced by merozoites and associated with immunity against malaria

## INTRODUCTION

The World Health Organization (WHO) recently recommended the widespread use of the RTS,S malaria vaccine in areas with moderate to high malaria transmission intensity^1^. Whilst a major landmark as no other malaria vaccine has advanced this far, RTS,S is modestly efficacious and the protection it induces wanes rapidly^2^. Vaccine development for malaria remains an urgent priority. Repeated exposure to *P. falciparum* parasitemia under conditions of high transmission intensity results in long term immunity to clinical episodes of malaria^3^. Passive transfer experiments demonstrated the importance of antibodies in mediating protection^4^, but the precise mechanisms that underpin their actions are still under investigation and there are no universally accepted correlates of protection. Numerous studies focus on the Fab-dependent neutralization of parasites that occurs for instance when antibodies inhibit the invasion of erythrocytes by merozoites, with inconsistent results^5^. Antibody-dependent Fc-mediated interactions with effector cells are increasingly being recognized as important correlates of protection^6–10^

The abundance of natural killer (NK) cells in peripheral blood positions them as prime immune effectors^11^. They are known to target malignant or virus-infected cells through an array of germ-line encoded activation receptors^12,13^. They can also be activated by opsonized malaria parasites via the low-affinity immunoglobin FcγRIIIa (CD16)^7^. Activation of NK cells leads to degranulation and the release of cytotoxic molecules like granzyme B, as well as the secretion of pro-inflammatory cytokines such as IFNγ^14^,with high Plasma levels of both associated with reduced parasitemia *in-vitro*^15^ and in cohort studies^16^. Recent studies have shown that malaria-immune sera target *P. falciparum* infected erythrocytes via NK cells leading to their destruction^9,17^. Merozoites may also be cleared in a similar fashion as antibodies to many merozoite antigens have been associated with protection against malaria and are considered as leading vaccine candidates^18,19^

We developed a novel antibody dependent natural killer cell assay (ab-NK) focused on *P. falciparum* merozoites. We demonstrate that anti-merozoite antibodies induce NK cell degranulation and IFNγ production in a largely strain-transcending fashion and enhance invasion-inhibition *in-vitro*. We use a human malaria challenge study in adults and an independent prospective cohort study in children and demonstrate for the first time that ab-NK targeting merozoites is predictive of protection. We identify specific merozoite antigens that induce ab-NK and provide a novel mechanistic correlate to support the prioritization of vaccine candidates.

## RESULTS

### Ab-NK activated following co-incubation with malaria-immune sera and merozoites

We tested whether merozoite-specific antibodies activated NK cells that were freshly isolated (immediately following blood draw) from malaria-naïve donors. We measured the levels of the classical surface marker of NK cell degranulation (CD107a) and intracellular IFNγ production. Activation was dependent on the presence of both malaria-immune sera and merozoites (Fig. 1A and B). Malaria-immune but not naïve sera activated NK cells in a dose-dependent fashion (Fig. S1A and B). We minimized assay variability by pooling NK cells from three independent donors in each experimental run (Fig. 1B). We validated the assay further using individual samples from malaria-exposed adults from Junju, Kenya, a region with moderate malaria transmission intensity (Fig. 1C & 1D).

**Figure 1.**
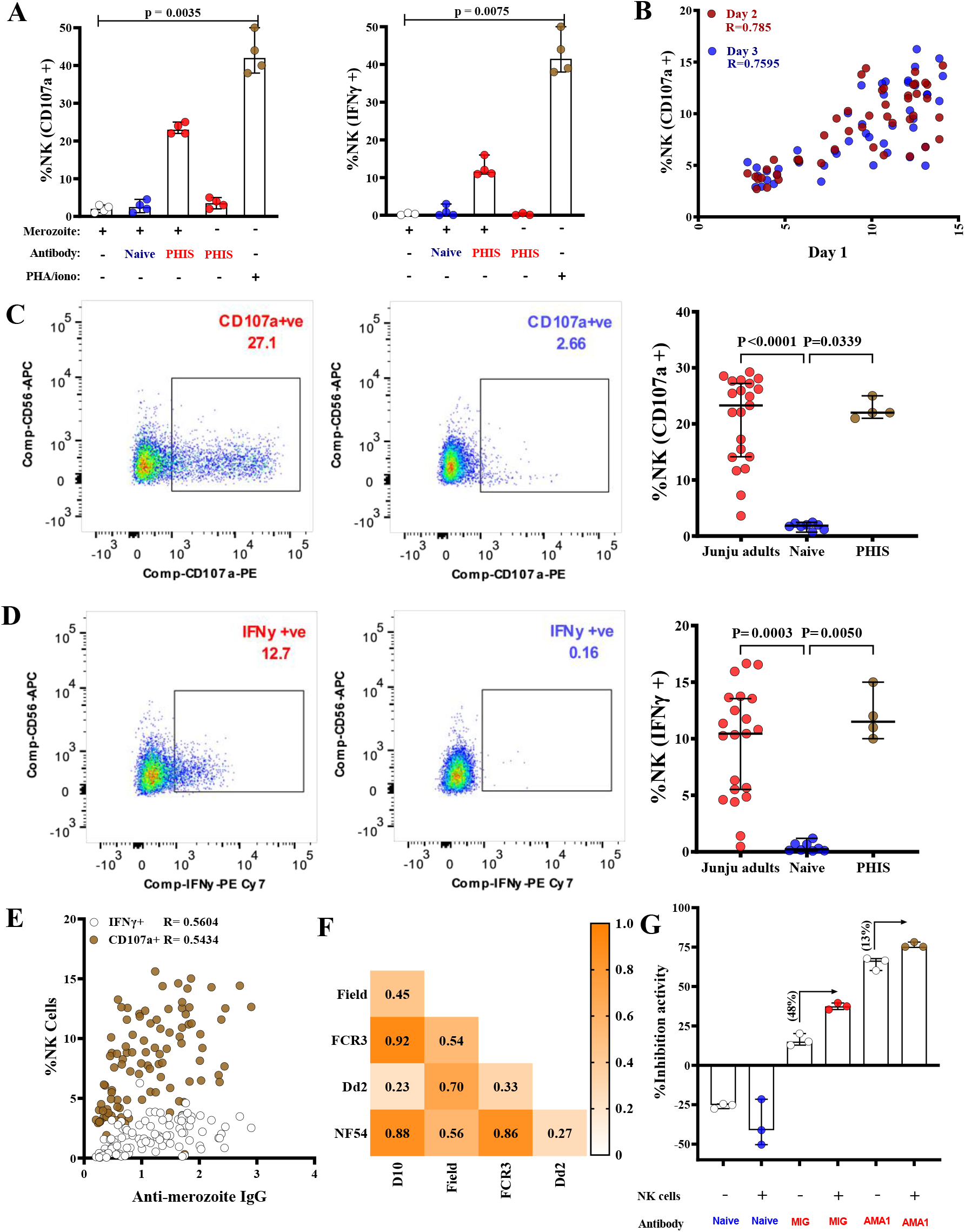
Anti-merozoite antibodies activate NK cells. **(A)** NK cell degranulation and IFNγ production was measured by CD107a surface and intracellular staining respectively. Merozoites were opsonized with a pool of malaria-immune or naïve plasma with PHA/ionomycin included as a positive control. Data are from four independent experiments and show the median with 95% confidence interval (95% CI); Kruskal-Wallis test. (**B**) Inter-assay variation was evaluated between days 1 versus 2 or 3. Pooled NK cells from 3 independent donors were tested using the same 30 malaria-immune samples on 3 separate days.(**C & D**) *P. falciparum* merozoites opsonized with Plasma from individual malaria-exposed adults (red circles, n=21) or malaria-naïve plasma (blue circles, n=4) were co-incubated with donor NK cells. Representative flow plots of NK cells incubated with merozoites in the presence of the respective plasma are shown. An additional pool of hyper immune sera (PHIS, comprised of sera from Junju adults with high ELISA responses against the 3D7 schizont lysate; brown circles, n=4) was included as a positive control. Error bars represent 95% CI of the median values; Kruskal-Wallis test. (**E**) Pairwise correlation between the proportion of IFNγ secreting or degranulating (CD107a+) NK cells activated by opsonized *P. falciparum* 3D7 merozoites and merozoite ELISA quantifying total IgG responses (n=142). (**F**) Spearman’s correlation heatmap between the proportion of NK cells degranulating upon activation by opsonized merozoites from five *P. falciparum* strains of different geographical origins (n=20). (**G**) Viable merozoites were co-incubated with uninfected erythrocytes and test immunoglobulins in the presence or absence of donor NK cells. Data represent the median with 95% confidence intervals of three independent experiments.

As expected, ab-NK was significantly correlated with total IgG against merozoites, Spearman’s R = 0.54 and 0.56 for CD107a and IFNγ respectively, P < 0.0001 (Fig. 1E). This correlation was greater for the cytophilic IgG1 and IgG3 isotypes (CD107a R = 0.54 & R = 0.51, P < 0.0001), when compared with the non-cytophilic IgG2 and IgG4 (CD107a R = 0.47 & R = 0.39, P < 0.0001; Fig. S2 A and B).)

### Ab-NK is strain-transcending across African isolates of *P. falciparum*

We compared ab-NK against five *P. falciparum* parasite isolates using 20 samples from malaria-exposed adults from Junju. Four of the five isolates were long-term laboratory-cultured strains of diverse geographical origin; NF54, FCR3, and D10 from West Africa and Dd2 from South East Asia. The fifth was isolated in Kenya (East Africa), has recently been adapted to laboratory culture conditions and is subsequently referred to as the clinical isolate. The pairwise correlation coefficients for ab-NK degranulation were significant and high between the laboratory isolates of African origin (NF54, FCR3, D10; Spearman’s R = 0.86 to 0.92, P < 0.001) but lower when compared to the clinical isolate (Spearman’s R = 0.45 to 0.56, P = 0.010, Fig. 1F). The latter may relate to the deletion of genes following long-term parasite culture^20^. The pairwise correlation coefficient between the SE Asian (Dd2) versus West African laboratory isolates (NF54, FCR3, D10) was markedly lower (Spearman’s R = 0.23 to 0.33, P = 0.245, Fig. 1F), consistent with the distinct genomic architecture observed between parasite isolates from Asia and Africa^21,22^. Surprisingly, a significantly higher correlation was observed between the Kenyan clinical isolate and the SE Asian laboratory strain (Spearman’s R = 0.70, P < 0.001) suggesting that these are more closely related. Similar results were observed for ab-NK IFNγ production levels (Fig. S3)

### Ab-NK enhanced *P. falciparum* invasion inhibition

*P. falciparum* merozoite invasion is a complex process that occurs within a narrow time window following schizont egress. To test whether ab-NK had an impact on merozoite invasion *in-vitro*, we used a modified standard invasion inhibition assay. Donor NK cells, uninfected red blood cells and freshly isolates merozoites were co-cultured in the presence or absence of antibodies. Malaria immunoglobulin (MIG) but not naïve antibodies inhibited parasite invasion. Interestingly, NK cells enhanced MIG-mediated invasion inhibition by 48%. Anti-rAMA-1 (polyclonal antibodies raised in immunized rabbits) was included as a positive control for invasion inhibition (Fig. 1G).

### Ab-NK is associated with protection following malaria challenge

We next assessed ab-NK in samples collected on the day before challenge (C-1) in a controlled human malaria infection (CHMI) study in semi-immune Kenyan adults^23^. The study endpoints were clinical symptoms of malaria with any evidence of malaria parasites by blood film positivity or parasitemia > 500 parasites/µl, both of which warranted immediate treatment^23^. Treated volunteers were further classified into those who developed fever and those who did not (febrile versus non-febrile). Untreated volunteers were sub-classified based on parasite detection by PCR into PCR-ve and PCR+ve groups^24^.

We asked whether ab-NK before challenge was associated with protection. Volunteers who were not treated (protected) had significantly higher levels of ab-NK cell degranulation and IFNγ than those that were treated (unprotected). (Mann-Whitney t test, P < 0.00001; Fig. 2A and 2B). Among the treated volunteers, there was no difference in ab-NK between those that were febrile versus non-febrile. Likewise, in the untreated (protected) sub-group, there was no difference between those who were either PCR + or PCR-ve. (Fig. 2C). To test for associations with protection, we stratified ab-NK cell into high versus low sub-groups based on threshold values derived from maximally selected rank statistics^25^. For both degranulation and IFNγ production, high ab-NK was associated with a significantly lower risk of treatment (incidence rate ratio 0.42 and 0.47 respectively, P < 0.001; Table 1). Furthermore, the time to detection of clinical malaria (study endpoints) after the tenth day post-challenge was longest in volunteers with high compared to low levels of ab-NK, Cox regression hazard ratio (HR) 0.26 and 0.19 for degranulation and IFNγ production respectively, P < 0.001; Table 1, Fig 2D). These analyses were adjusted for the potential confounding effects of low levels of lumefantrine (below the minimum inhibitory concentrations) that were detected in the C-1 samples, and the geographical origin (location of residence) of the volunteers.

**Table 1:**
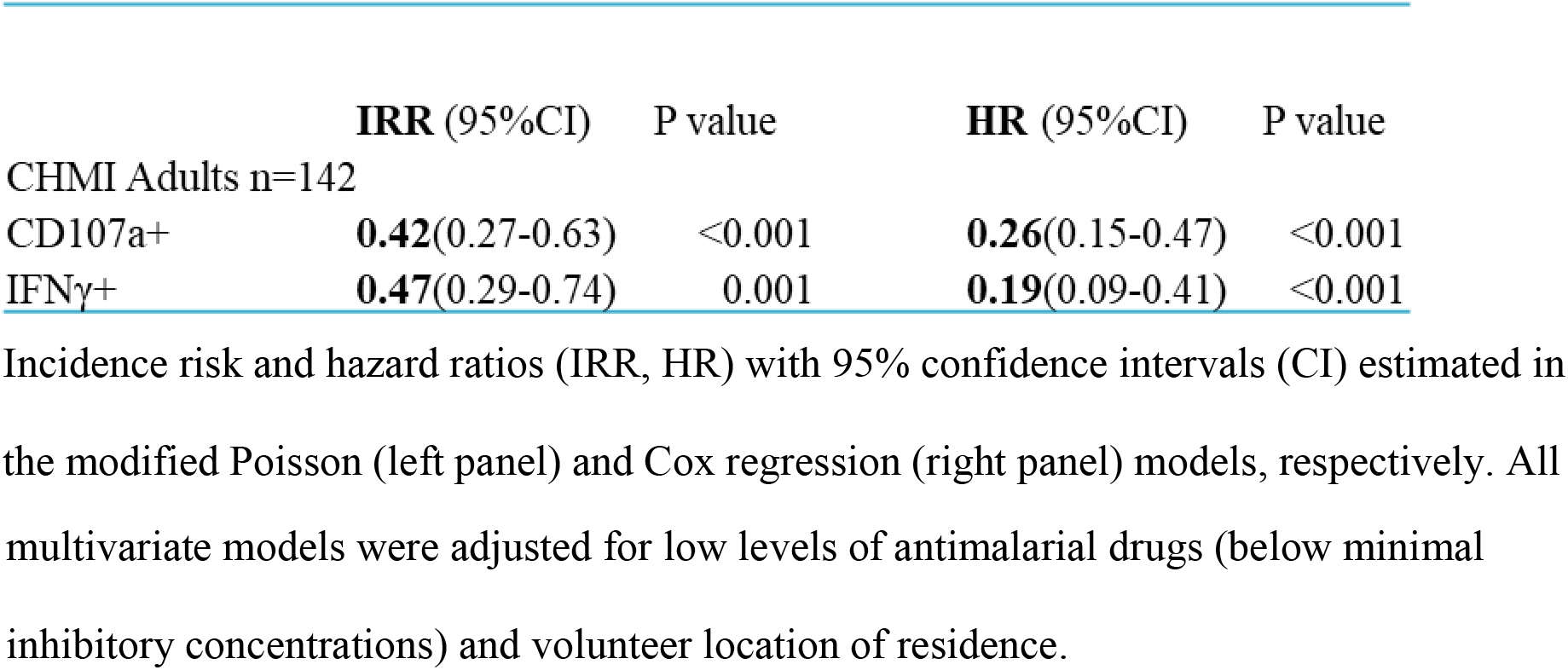
Ab-NK predicts protection following malaria challenge.

|  | **IRR (95%CI)** | **P value** | **HR (95%CI)** | **P value** |
| --- | --- | --- | --- | --- |
| CHMI Adults n=142 |  |  |  |  |
| CD107a+ | **0.42(0.27-0.63)** | <0.001 | **0.26(0.15-0.47)** | <0.001 |
| IFNγ+ | **0.47(0.29-0.74)** | 0.001 | **0.19(0.09-0.41)** | <0.001 |

Incidence risk and hazard ratios (IRR, HR) with 95% confidence intervals (CI) estimated in the modified Poisson (left panel) and Cox regression (right panel) models, respectively. All
multivariate models were adjusted for low levels of antimalarial drugs (below minimal inhibitory concentrations) and volunteer location of residence.

**Figure 2.**
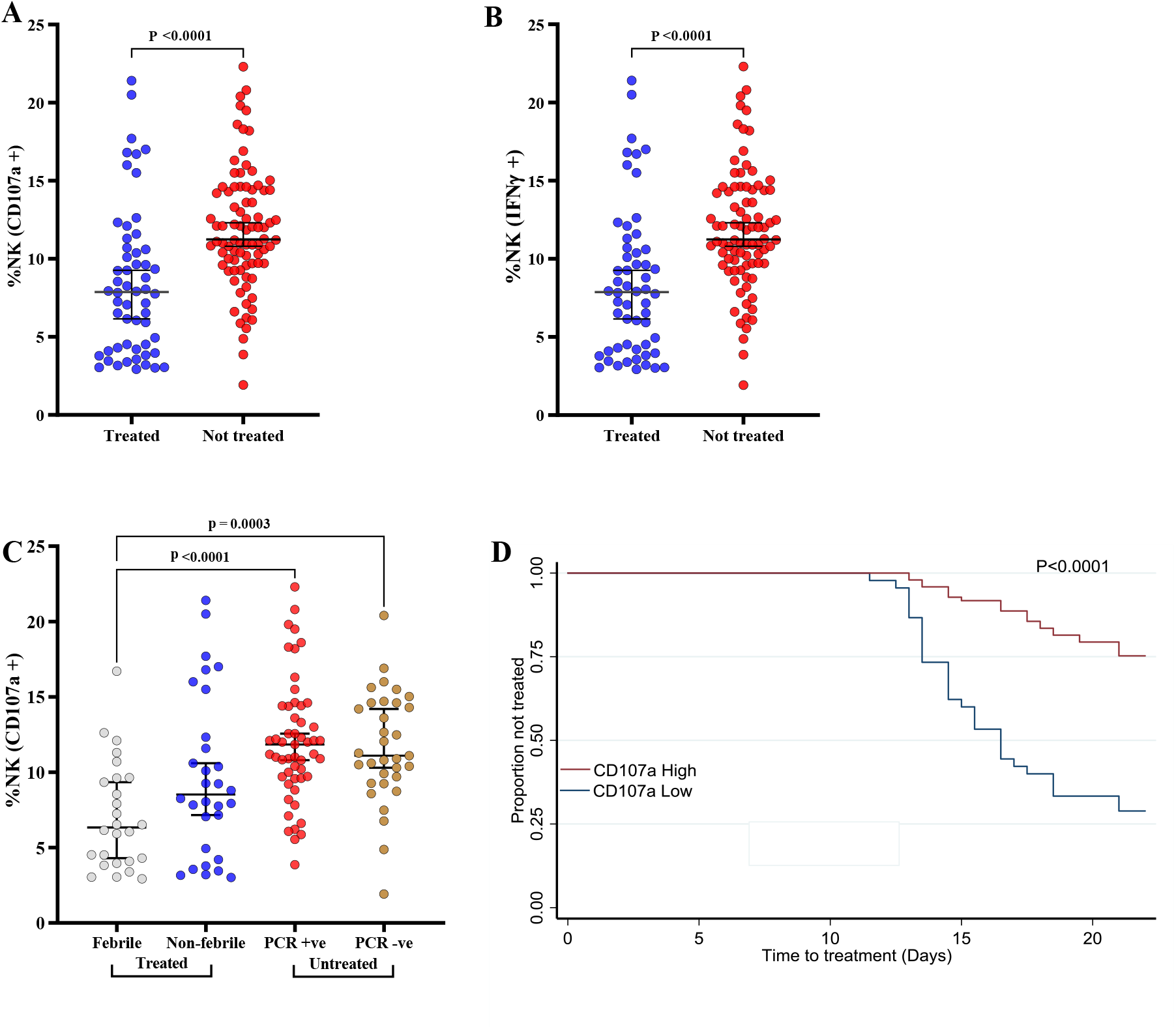
Antibody-mediated NK cell activity is associated with *in vivo* parasite growth. Comparison of ab-NK degranulation (**A**) and IFNγ production (**B**) in treated (n=56) versus non-treated (n=86) volunteers. Bars represent 95% CI of the median values; Mann-Whitney test. (**C**) Sub-group analysis ab-NK degranulation for treated volunteers who either developed fever (febrile n= 26) or did not (non-febrile n=30), and for untreated volunteers in whom parasites were either detected by PCR (PCR+ve n=53) or remained negative (PCR-ve n=33). Bars represent 95% CI of the median values; Kruskal-Wallis with Dunn’s multiple comparison test. (**D**) Kaplan-Meier curves for the time to treatment for volunteers with a high (red) versus low(blue) ab-NK degranulation; log-rank test, P<0.0001 n=142.

### Not all potential merozoite vaccine candidate antigens induce Ab-NK

To distinguish distinct merozoite antigens that induce ab-NK from those that potentially did not, we developed a plate-based ab-NK assay utilizing recombinant antigens. We could thus detect and quantify antigen-specific ab-NK. We tested ab-NK for 14 *P. falciparum* merozoite surface-associated antigens identified as potential vaccine candidates in a previous study^18^. This analysis was conducted in a subset of CHMI samples with high (n=8) or low (n=2) levels of ab-NK against whole merozoite extract. We found that antigen-specific ab-NK varied between antigens and individuals. We identified 8 potential merozoite targets of ab-NK out of the 14 tested (Fig. 3A). These included well-characterized vaccine candidates like AMA-1, MSP3 and MSP2 and less-well studied antigens like P41, P113, MSP11, RHOPH3, and *Pf*_11363200 that have independently been associated with clinical protection^14–16^. (Fig. 3A). Responses against other leading blood stage vaccine candidates such as *Pf*Rh5 and EBA-175 had negligible ab-NK. Although we had pre-selected individuals with high ab-NK degranulation against whole merozoite extract, this did not always translate into high levels of antigen-specific ab-NK. This suggests that additional antigens that were not tested here contributed to the functional response against the whole merozoite (Fig. 3B). Additionally, we and others previously found that the breadth of the antibody response was associated with enhanced function^19,28,29^. In keeping with this, whole merozoite extract ab-NK cell degranulation and the breadth of antigen-specific ab-NK cell degranulation were strongly correlated (Fig. 3C and Fig 3D).

**Figure 3.**
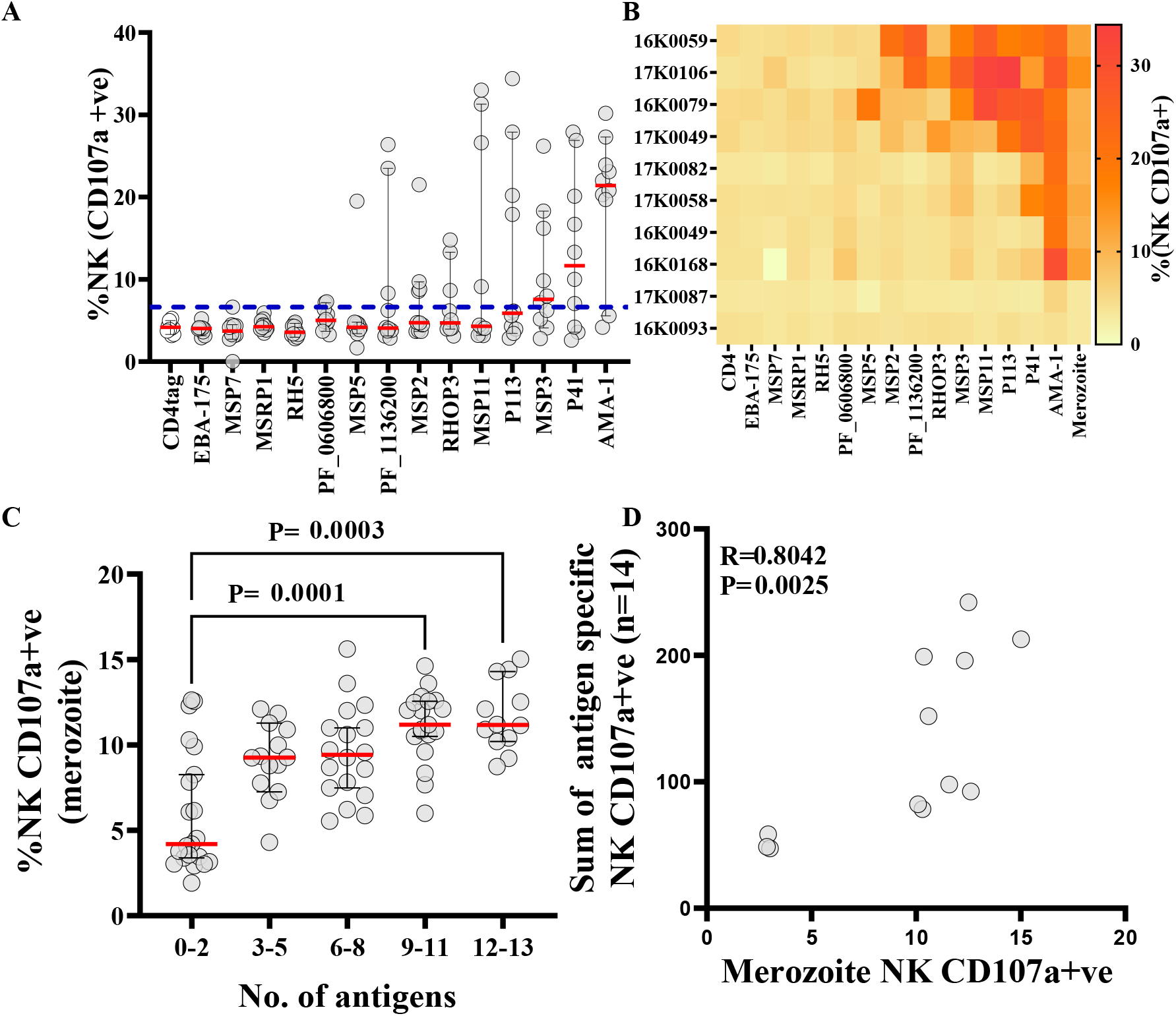
Potential targets of antibodies that mediate ab-NK. (**A**) Antigen-specific ab-NK cell degranulation from a subset of CHMI individuals (n=10) against 14 unique recombinant *P. falciparum* merozoite antigens. Blue line represents the cut-off value based on the mean plus 3 standard deviations of the tag fragment (CD4 tag). Error bars represents 95% CI of the median (Red line). (**B**) Heatmap of individual level antigen-specific ab-NK cell degranulation from A. Each row represents a single CHMI participant and each column represents responses to a single recombinant merozoite surface associated antigen. (**C**) Ab-NK cell degranulation against whole merozoite extract was associated with the breadth of antigen-specific recognition. Error bars represents 95% CI of the median (Red line) n= 142; Kruskal-Wallis test. (**D**) The sum of antigen-specific ab-NK cell degranulation against 14 unique recombinant merozoite antigens was correlated with NK cell degranulation against merozoites; Spearman correlation R = 0.80.

### Ab-NK increases with age following natural infections

We next examined the acquisition of antibodies that mediated NK cell activity in 293 samples collected from a longitudinal cohort study of children living in Junju, Kenya^30^. At the time of sampling, 67% of the children had antibodies that mediated NK cell degranulation while only 10% had antibodies that induced IFNγ production, respectively (Table 2). Notably, we observed significantly lower ab-NK in children compared to adults living in the same location (Fig. S4A). Ab-NK increased with age (Fig. 4A; Fig. S4B). Additionally, children who were parasite slide positive at the time of sampling showed higher NK cell degranulation but not IFNγ production than slide negative children, suggesting a boosting effect of ab-NK by active *P. falciparum* infection (Fig. 4B). Antibody responses against three malaria vaccine candidates (AMA-1, MSP2 and MSP3) had been measured in this cohort in a previous study^30^. Ab-NK in children was modestly correlated with antibody recognition of AMA1 (CD107a+: R = 0.35 and IFNγ+: R = 0.31), MSP2 (CD107a+: R = 0.28 and IFNγ+: R = 0.35), and MSP3 (CD107a+: R = 0.25 and IFNγ: R= 0.24; Spearman’s R for all comparisons P< 0.0001; Fig. S4C). However, despite the modest correlation, the breadth of antibody recognition against the three merozoite antigens was associated with increased NK cell activity against the whole merozoite in children, again suggesting an additive effect when multiple targets are considered (Fig. 4C).

**Table 2:**
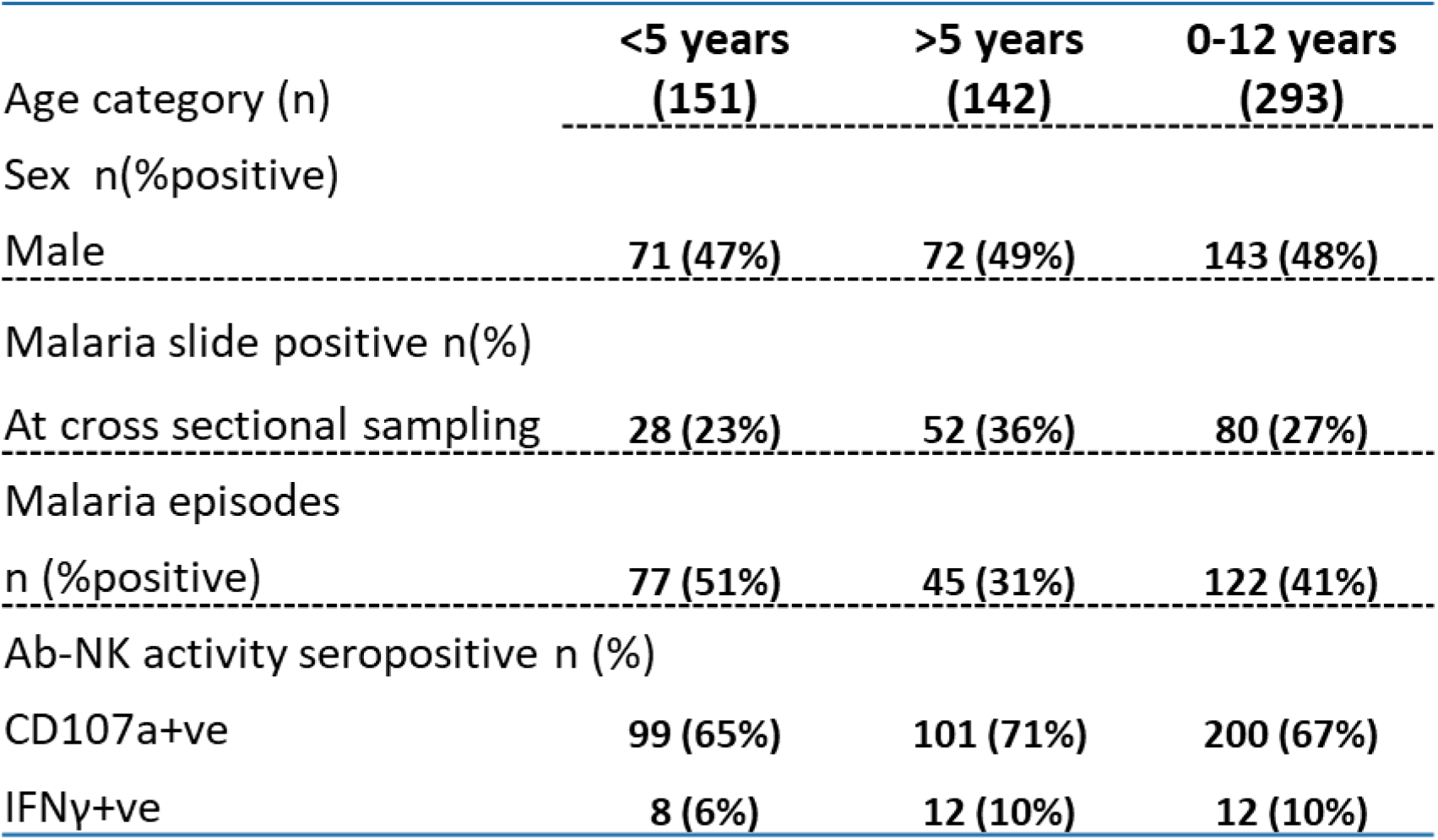
Junju cohort baseline characteristics.

|  | <5 years<br>(151) | >5 years<br>(142) | 0-12 years<br>(293) |
| --- | --- | --- | --- |
| Age category (n) |  |  |  |
| Sex n(%positive) |  |  |  |
| Male | 71 (47%) | 72 (49%) | 143 (48%) |
| Malaria slide positive n(%) |  |  |  |
| At cross sectional sampling | 28 (23%) | 52 (36%) | 80 (27%) |
| Malaria episodes<br>n (%positive) | 77 (51%) | 45 (31%) | 122 (41%) |
| Ab-NK activity seropositive n (%) |  |  |  |
| CD107a+ve | 99 (65%) | 101 (71%) | 200 (67%) |
| IFNγ+ve | 8 (6%) | 12 (10%) | 12 (10%) |

**Figure 4.**
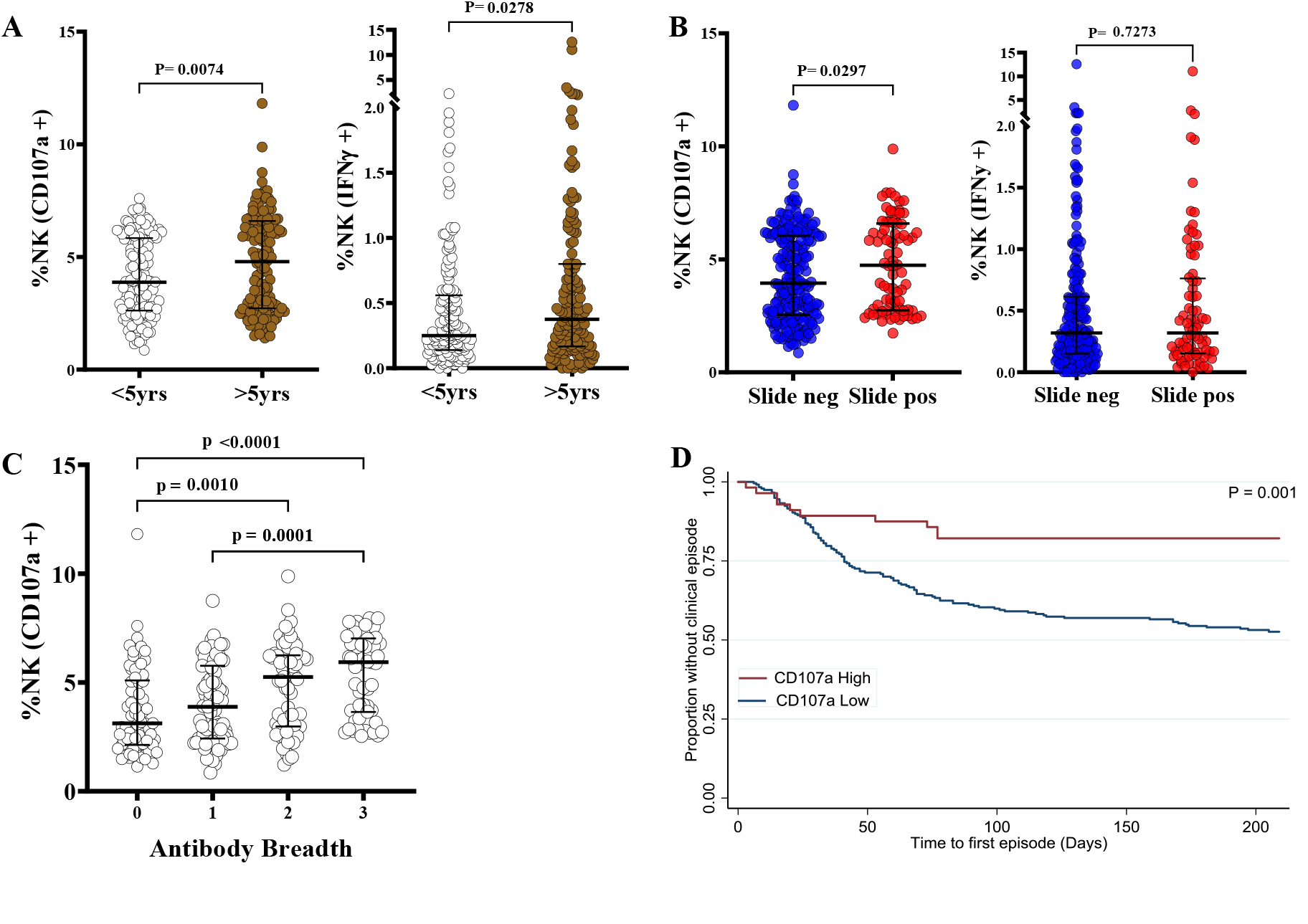
Antibody-mediated NK cell degranulation is associated with clinical protection in a prospective cohort study in Kenyan children. (**A**) Comparison of Ab-NK in Junju children over or under 5 years of age. (**B**) Ab-NK response in children who had an active *P. falciparum* infection compared to those without. Error bars represent 95% CI of the median values; Mann-Whitney test (n=293). (**C**) Antibody-mediated NK cell degranulation is correlated with the breadth of total IgG responses against recombinant MSP2, MSP3 and/or AMA-1. Error bars represent 95% CI of the median values; Kruskal-Wallis test with Dunn’s multiple comparison test (n=293). (**D**) Ab-NK cell degranulation is associated with clinical protection in children. NK cell degranulation responses were categorized as high or low (blue) based on a threshold^25^; log-rank test, P= 0.001 (n=293). Each dot represents a technical replicate.

### Ab-NK is associated with protection against clinical episodes of malaria in children

To date, no longitudinal cohort study has assessed the relationship between ab-NK against merozoites and protective immunity in children living in Kenya. We stratified ab-NK into high versus low categories based on derived thresholds^25^ and fitted them to a Poisson regression analysis, adjusting for age and previous *P. falciparum* exposure as confounders^6^. We found that both degranulation and IFNγ production were significantly associated with clinical protection CD107a (IRR: 0.34 (95% CI, 0.19-0.61); P < 0.000 and IFNγ; (IRR: 0.61 (95% CI, 0.38-0.97); P < 0.040; Table 3). These findings were confirmed in a Cox regression analysis in which the time to the first malaria episode was analyzed as the outcome variable (log-rank test, P<0.0001; Fig. 4D).

**Table 3:** Ab-NK is associated with a reduced risk of clinical malaria episodes in children from Junju, Kenya.

| **Junju Children**<br>0-12 years (n=293) | **IRR (95%CI)** | **P value** | **HR (95%CI)** | **P value** |
| --- | --- | --- | --- | --- |
| CD107a+ | 0.34(0.19-0.61) | 0.000 | 0.26(0.13-0.52) | 0.000 |
| IFNγ+ | 0.61(0.38-0.97) | 0.038 | 0.54(0.30-0.97) | 0.040 |

Ab-NK cell degranulation and IFNγ production were fitted to modified Poisson regression models. Results are presented as incidence risk ratios (IRR) or hazard ratios (HR) with 95% confidence interval (CI). P-values >0.05 were considered significant. All models were adjusted for age and previous *P. falciparum* exposure as confounders.

## DISCUSSION

Defining the mechanisms that underpin the potent efficacy of malaria immunoglobulin observed in passive transfer experiments provides a strong evidence-base for rational vaccine development. We report for the first time that anti-merozoite antibodies engage Fc receptors on natural killer cells, thereby unleashing potent anti-parasite effector activity. Ab-NK was associated with clinical protection in two independent studies and induced responses against multiple parasite strains, including a recently adapted clinical isolate. Additionally, we identify a subset of merozoite antigens that induce ab-NK and show that the breadth of antigen-specific ab-NK mirrors that quantified against merozoites. Our data augment the accumulating body of evidence that targeting combinations of antigens may be instrumental for malaria vaccine design^18,19,29,31–35^. We also provide a new mechanism for the prioritization and subsequent evaluation of vaccine candidates.

Although several immune mechanisms have been proposed as correlates of protection, none have focused on Fc-antibody-dependent targeting of merozoites by natural killer cells. The growth inhibition assay (GIA) is considered the gold standard for the assessment of antibody function and is thought to assess a combination of invasion-inhibition, growth inhibition and possibly merozoite egress^36–39^. Unfortunately, it does not reliably predict protection. Other studies have investigated the role of complement^7^, monocytes^6^ and neutrophils^8^, marking the increasing importance of Fc-mediated function^10^, with promising results that need further validation in additional studies Importantly, our unique challenge study allowed us to overcome many of the limitations of cohort studies that often lead to conflicting results when any functional correlate of protection is considered. The timing and intensity of our experimental parasite challenge was accurately defined, and close monitoring for clinical symptoms and parasite multiplication rates was feasible because volunteers were hosted at a single location for the duration of the study. We included adults from a single East African country to minimize genetic variability and pre-selected individuals with a varied range of previous malaria exposure to allow us to explore its’ impact on the clinical outcome. Under these stringent conditions, we observed a strong and significant correlation between ab-NK and protection. We found similar results in an independent study, involving only children, and that utilized an entirely different design in which malaria infections occurred naturally under real-life conditions.

Natural killer cells have been extensively studied for their role in the early defense against viral infections and cancers. Additionally, the modulation of their function underpins an array of contemporary immunotherapeutic agents^40,41^. Our study capitalizes on the unique ability of the FcγRIIIa (CD16) to induce NK cell in response to antibody-coated targets, without engaging other activating or inhibitory receptors^42^. The potential impact of this mechanism on vaccines against infectious diseases is relatively understudied. Recently, a human monoclonal antibody (mAb) against a conserved region of the haemagglutinin (HA) protein, was shown to not only potently neutralize a broad range of influenza viruses, but also to mediate ADCC^43^. In HIV, evidence that ADCC-mediating antibodies complement neutralizing functions is accumulating^44,45^.

In malaria, natural killer cells were identified as part of the early innate immune response, through the production of IFNγ and soluble granzymes^14^. Although recent data suggest that they can acquire a memory-like “adaptive” phenotype following repeated exposure with malaria^17,46^, our study utilized malaria-naïve NK cells to interrogate their interaction with anti-merozoite antibodies. As shown for malaria antigens exposed on the surface of infected red cells^9,46,47^ antibodies against merozoites induce Fc-receptor dependent NK activation. In our studies, antibodies inducing ab-NK increased with age, were boosted by recent malaria infections and were strongly predictive of immunity. Furthermore, our antigen-specific ab-NK assay provides novel insights on individual merozoite antigens that induce functional antibodies and could guide rational vaccine design.

The detection of functional activity against a diverse panel of parasite strains is noteworthy and a vitally important consideration when new assays and vaccine candidates are assessed^5,35^. Previous studies investigating antibody-dependent NK effector function in *P. falciparum* focused on a single laboratory strain^9,17,46,47^. We found that ab-NK responses against merozoites from diverse *P. falciparum* strains originating in Africa were strongly correlated. Although this suggests that some of these antibodies target conserved epitopes, further confirmation at the antigen and indeed epitope level is needed. Interestingly, the weaker correlations between the African laboratory versus clinical isolate indicate that some antibodies do target polymorphic epitopes. In influenza, ADCC-mediating antibodies target conserved epitopes in the HA protein^48^, raising the exciting possibility of broadly protective universal influenza vaccines.

We developed a high-throughput plate-based antigen-specific ab-NK cell assay^49^ to home-in on specific merozoite antigens targeted by ab-NK. We tested a pragmatic panel of merozoite antigens that were available to us from previous studies in a subset of samples from adults^50^. We observed high inter-individual variation and unique ab-NK antigen-specific reactivity profiles in each sample. This was not surprising given the heterogeneity of antibody responses against malaria antigens that we, and others, have previously reported^18,51^. Interestingly, leading vaccine antigens known to induce invasion-inhibitory antibodies such as AMA1^52^, or Fc-dependent activity in phagocytosis such as MSP3^6^, were also targeted by ab-NK in some, but not all individuals. Less-well studied vaccine candidates such as *Pf*113 and P41 also induced ab-NK in a proportion of individuals. Notably, some individuals elicited high levels of ab-NK against the whole merozoite, but not against a broad range of tested antigens. Further dissection of antigen and epitope specificity is warranted and may be instructive for vaccine design.

Previous studies have demonstrated that the breadth of the antibody response against selected parasite antigens is an important predictor of protection against malaria^18,19,29,31–35,53^. The breadth of antibody reactivity against the 14 *P. falciparum* antigens tested in the current study was associated with higher ab-NK against the whole merozoite. This mirrors observations from other functional studies in which the breadth of the antigen-specific functional Fc-antibody dependent response was correlated with protection^18,19^. These data suggest that an effective malaria vaccine modelled on naturally acquired immunity may not only need to incorporate multiple antigens, but also induce antibodies that trigger diverse effector functional activity. The influenza example is a case in point; a multifunctional human mAb was identified that simultaneously neutralizes virus and induces ADCC^43^.

We identify antibody-mediated NK cell activity targeting merozoites as a strong predictor of naturally acquired immunity against *P. falciparum* malaria. Our antigen-specific assay facilitates throughput and will galvanize additional studies focusing on specific vaccine candidates. Further dissection of ab-NK inducing antibodies at the epitope level may reveal important signatures for universal vaccine design.

## MATERIALS AND METHODS

### Study design and samples

#### Controlled human malaria infection (CHMI) study

The study design and details of the CHMI study have been described previously^54^. Briefly, a dose of 3200 infectious, cryopreserved *P. falciparum* NF54 sporozoites (Sanaria *Pf*SPZ) was administered intravenously to 161 consenting Kenyan adults (18-45 years) with varying degrees of prior exposure to malaria (based on ELISA responses to crude *P. falciparum* 3D7 schizont lysate). Testing for parasitemia was conducted twice daily from day 7 to 14; and once from day 15 to 21 days post-challenge by qPCR. Subjects were treated either when they had more than 500 *Pf* /μL, exhibited clinical symptoms and had any parasitemia or at the end of the active follow up period on day 21. Data from 19 individuals were excluded due to detectable drug levels^54^ (Fig. S6). We analyzed 142 plasma samples collected one day before the challenge (C-1) to test whether ab-NK was associated with a lower risk of clinical malaria or parasitemia > 500 *Pf* /μL.

#### Longitudinal study: Junju cohort

The Junju cohort was established in 2005 as part of a malaria vaccine trial^55^, and participants have been followed up since then. Briefly, we analyzed 304 plasma sample collected at the beginning of the malaria transmission season in May 2008 from children aged between 0-12 years and resident in Junju village, Kilifi County. The *Plasmodium falciparum* parasite rate standardized to the age group 2–10 years (*Pf*PR_2–10_) was 29% at the time of sampling. Active follow-up involved weekly visits to participants homes where a questionnaire for clinical symptoms was administered and temperature was recorded. In addition, study personnel were resident in the village and were contacted by caregivers contacted at any time if the children were unwell. A clinical malaria episode was defined either as a temperature of >37.5°C plus any parasitemia for children under 1 year, or a temperature of >37.5°C plus a parasitemia of >2500 *Pf* /μL for children older than 1 year^56^. We analyzed data spanning the malaria transmission season of approximately 6 months duration. Independent samples collected from adults in Junju (n = 40) were used to establish and optimize the ab-NK cell assay. A pool of hyper-immune sera collected from malaria-exposed adults (PHIS) from Kilifi, Kenya and a commercial Malaria Immune Globulin^57^ (MIG) were used for validation experiments and as positive controls.

### Cultivation, purification and quantification of viable *P. falciparum* merozoites

*P. falciparum* laboratory adapted strains of West African (NF54, FCR3, D10) and SE Asian origin (Dd2), as well as a recently adapted clinical isolate from Kilifi, Kenya (P0000072) were cultured in RPMI-1640 media supplemented with 0.5% Albumax, 25µg/mL gentamycin, 50µg/mL Hypoxanthine, 2mM L-glutamine and 25mM HEPES buffer. Cultures were maintained at 2% parasitemia with O-positive erythrocytes obtained from malaria-naïve donors (less than 2 weeks old). Free viable merozoites were isolated as previously described ^58^.Briefly, *P. falciparum* cultures were allowed to attain 10-15% parasitemia and synchronized at the ring stage using 5% sorbitol. After 24 hours, late stage pigmented trophozoites were harvested by Magnetic Activated Cell Sorting (MACS) column purification as per the manufacturer’s instructions (Miltenyibiotec), attaining trophozoite purifications of 80-95%. The enriched trophozoites were put back in culture and allowed to develop until they began to undergo schizogony (segmented nuclei) whereby 1mM of a protease inhibitor (E64; SIGMA) was added to allow maturation into late schizonts but inhibit rupture. Mature schizonts were collected and passed through a 1.2μm microfilter previously blocked in 1% casein in PBS for 10 minutes to release free viable merozoites. These were either immediately used in the IIA-ADCC assay or washed twice in PBS, quantified and stored at - 80°C until needed. The relative concentration of the stored merozoites was determined using CountBright(tm) absolute counting beads as per the manufacturer’s instructions. In brief, 50µL of counting beads with a known concentration was mixed with a known volume of merozoites stained with 100μL 1X SYBR® dye for 15 minutes at 24°C and acquired on a BD FACSCaliburII flow cytometer. Merozoites were resuspended in 1XPBS at a working concentration of 5×10^7^ merozoites/mL and stored at -80°C.

### Recombinant expression of *P. falciparum* merozoite antigens

The recombinant merozoite antigens were expressed in-house as previously described^50^. In brief, Expi293F cells (Invitrogen) were cultured to a density of 2.0 × 106 cells/mL and transfected with expression vectors using the Expifectamine 293 transfection reagent (Invitrogen). Cells were then incubated at 37°C with 8% CO2 in an orbital shaker at 125 rpm. Culture supernatants were harvested 6 days post-transfection and proteins purified using Ni-NTA purification columns (Invitrogen).

### ELISA

A standardized ELISA protocol was performed as published^6,29,30,59^. In brief, a pre-determined concentration of recombinant *P. falciparum* recombinant antigens (30ng/well) or free whole merozoites (5×10^5^ merozoites/well) were coated overnight at 4°C. The plates were washed with PBS followed by blocking with 1% casein in PBS (Invitrogen) for 2 hours at 37°C. After four washes, 50μL of test plasma sample diluted 1:20 in PBS was added to the plates in duplicate and incubated for 1 hour at 37°C, followed by four additional washes. The plates were incubated for 1 hour at 37°C with 100μL of secondary anti-human IgG, IgG1, IgG2, IgG3 or IgG4 (Invitrogen) antibodies used to determine total IgG or IgG isotype specific antibody responses, respectively. Plates were washed and 100μL of OPD substrate (Invitrogen) in PBS was added and incubated for 20 minutes for color development. The reaction was stopped with 1M hydrochloric acid solution (Sigma) and optical density quantified at 492nM. Plasma samples from a pool of hyper immune adults (PHIS) and a pool of malaria-naïve German donors were used as positive and negative controls respectively. Samples were considered seropositive if they had an optical density higher than the mean plus 3 standard deviation of the naïve controls.

### NK cell isolation

Venous blood was collected from healthy malaria-naïve donors and PBMCs were subsequently isolated within 4 hours of collection using density gradient separation on a histopaque (1077g/dL) monolayer. The PBMCs were washed twice in PBS, assessed for viability using trypan blue exclusion and resuspended in cold RPMI-1640 media supplemented with 10% fetal calf serum (Sigma) at a final concentration of 1.0×10^7^cells/mL. The NK cells were isolated from the PBMCs by depleting non-NK cells lymphocytes (negative selection) using an NK isolation kit as per the manufacturer’s instructions (Miltenyibiotec). Briefly, 20µL of a cocktail of monoclonal antibodies was added to 1.0×10^7^ PBMCs and incubated for 30 minutes followed by 30 minutes incubation with 40µL of microbeads at 4°C. The PBMC mixture was then passed through a MACs column followed by 5ml of cold RPMI-1640 before collecting NK cells in the flow-through. The enriched NK cell mixture was washed, resuspended in fresh NK cell medium, (RPMI-1640, 10% FCS, 1% Penicillin/streptomycin and 2mM L-glutamine) and used on the same day. The NK cell isolation efficiency and cell purity (> 90%) was confirmed by flow cytometry (Fig. S1D).

### NK cell degranulation and intracellular cytokine assay

Whole merozoites (4.5×10^6^ merozoites/well) or recombinant *P. falciparum* antigens (30ng/mL) were coated overnight in 96 well-culture plates at 4°C. The wells were washed with PBS and blocked for 2 hours at 37°C with 1% casein in PBS. The coated plates were opsonized with heat-inactivated, pre-diluted (1:20) plasma samples for 5 hours at 37°C. Freshly isolated NK cells were added into each well (2.0×10^3^ NK cell/well). Anti-human CD107a PE (1:50), brefeldin A and monensin (5μg/mL) were added and the plate was incubated for 18 hours at 37 °C in 5% CO^2^. The NK cells were washed, centrifuged at 1500 rpm for 5 minutes and resuspended in FACS buffer (PBS, 1% BSA, 1mM EDTA and 0.1% sodium azide). Their viability was assessed by staining with a fixable viability dye eFluor ^™^520 for 10 minutes at 4°C. The temperature was maintained at 4°C for all subsequent steps. A cocktail of anti-human monoclonal antibodies comprising of anti-CD56 APC, anti-CD3 PE-Cy5 and anti-CD16 APC-Cy7, was used to stain the corresponding NK cell surface markers for 30 minutes. The cells were subsequently washed twice with FACS buffer; fixed with CELL fix (BD) for 10 minutes and permeabilized with a permeabilization buffer (BD) for 10 minutes. The permeabilized NK cells were stained intracellularly with anti-human IFNγ PE-Cy7 for 1 hour in the dark. Finally, the NK cells were washed thrice in permeabilization buffer, resuspended in FACS buffer and stored at 4°C awaiting acquisition. Controls wells included: 1) un-opsonized merozoites, 2) merozoites opsonized with malaria-naïve plasma, 3) merozoites opsonized with PHIS, 4) merozoites incubated with PBS only, and 5) NK cells incubated with 1ng/mL of Phytohemagglutinin (PHA) and 1ug/mL ionomycin (nonspecific NK cell stimulants). Acquisition was performed on the BD FACSCaliburII high-throughput system in a 96-well plate format using FACSDiva®. Data analysis was performed using Flow.Jo V10.

### Modified invasion inhibition assay (IIA-ADCC)

Approximately 20,000 NK cells/well, uninfected erythrocytes and antibodies from malaria-naïve or malaria-exposed adults were resuspended in *P. falciparum* culture media at a final hematocrit of 0.5%/well) in a 96-well plate format. Synchronized schizonts (500µL pellet) from *P. falciparum* strain NF54 was resuspended in 2500µL *P. falciparum* culture media and filtered through a 1.2µM membrane to release viable merozoites^58^ and immediately added into each well (40µL/well). The culture plate was incubated in a shaking incubator (50 rpm) for 30 minutes at 37°C to promote invasion. Purified polyclonal anti*-*rAMA-1(1.5ng/well), pooled malaria-naïve immunoglobulin (1.5ng/well) and a commercial anti-malaria immunoglobulin, MIG (1.5ng/well) were tested. After the 30 minutes co-incubation of merozoites, antibodies and NK cells; unbound antibodies and free merozoites were removed by washing the culture twice in fresh *P. falciparum* culture media. Subsequently, NK cells were removed using density gradient separation by gently adding 80µL/well of histopaque (1077g/dl) and spinning at 1800 rpm for 15 minutes without brakes. The culture was then washed twice, resuspended in *P. falciparum* culture media (100µL/well) and maintained for an additional 30 hours at 37°C. At the end of incubation, parasites were stained using the SYBR dye and enumerated by flow cytometry. Invasion inhibition was calculated against the proportion of the parasitemia recorded in the reference wells (test well without any antibody or NK cells)

### Flow analysis

Flow data was analyzed using FlowJo V10.1 (TreeStar). For the analysis of phenotypic data, positive gates for CD56, CD16 and CD3 were used to define NK cells. For the antibody mediated functional data, positive gates for intracellular cytokine production and degranulation were included with the resulting proportion of each subset tabulated.

### Statistical Analysis

Data was analyzed using Prism 8.07 (GraphPad) or Stata (version 14). The Mann Whitney U test was used to compare medians between distinct pairs. The Kruskal-Wallis test was use to compare more than two groups and supplemented by Dunn’s test for multiple comparisons. A nonparametric Spearmans’ correlation was used to estimate the strength of pairwise correlations. The threshold level (analytical cutoff) above which ab-NK was associated with protection was derived using maximally selected rank statistics^25^. The responses were grouped into two groups (high and low). Associations with protection were assessed in both studies using the modified Poisson and Cox regression models^6,29,30^. Potential confounders were adjusted to the respective models and included; detectable levels of lumefantrine in the sample collected one day prior to challenge and the location of residence.in the CHMI study. For the Junju cohort, we adjusted for, age and schizont reactivity as a proxy for previous exposure.

## Data Availability

The study protocol and outcomes are published23. Additional original data that support the findings of this study are available from the data governance committee at KWTRP upon reasonable request;

## Supplementary Materials

Fig. S1. Optimization of novel ab-NK assay

Fig. S2. Ab-NK activity is correlated with anti-merozoite antibody binding

Fig. S3. Ab-NK activity mediates strain transcending responses

Fig. S4. Antibody mediated NK cell activity in children.

Fig. S5. Flow cytometry gating strategy

Members of the CHMI-SIKA study team.

## Acknowledgments

We thank all the study volunteers who participated in the CHMI-SIKA and Junju study. We are thankful to the larger study teams in Kilifi responsible for recruitment, data entry, sample processing and storage. Additionally, we appreciate all our malaria-naïve donors who provided us with fresh PBMCs.

## Author contributions

DOO and FHAO conceived and designed the experiments. DOO, FHAO, RF and KM performed the analysis. JT, TC and RC contributed recombinant antigens. DOO, INN, KF, SD and MR performed experiments. FHAO, PN, MH, MCK, provided the clinical samples. DOO, RF and FHAO wrote and edited the paper. All authors contributed to revision of the manuscript.

## Ethical Statement

The CHMI study was conducted at the KEMRI Wellcome Trust Research Programme in Kilifi, Kenya with ethical approval from the KEMRI Scientific and Ethics Review Unit (KEMRI//SERU/CGMR-C/029/3190) and the University of Oxford Tropical Research Ethics Committee (OxTREC 2-16). All participants gave written informed consent. The study was registered on ClinicalTrials.gov (NCT02739763), conducted based on good clinical practice (GCP), and under the principles of the Declaration of Helsinki.

The Junju cohort, were originally recruited in 2005 and has been followed up, for clinical episodes of malaria^55^. In this study we used samples collected in 2008. Ethical approval for the Junju study was provided by the Kenyan National and Scientific Ethics Review Committee protocol number 3149.

## Data and material availability

The study protocol and outcomes are published^23^. Additional original data that support the findings of this study are available from the data governance committee at KWTRP upon reasonable request;

