## Supplementary data for "Antibodies targeting merozoites induce natural killer cell degranulation and interferon gamma secretion and are associated with immunity against malaria"

### Supplementary Materials

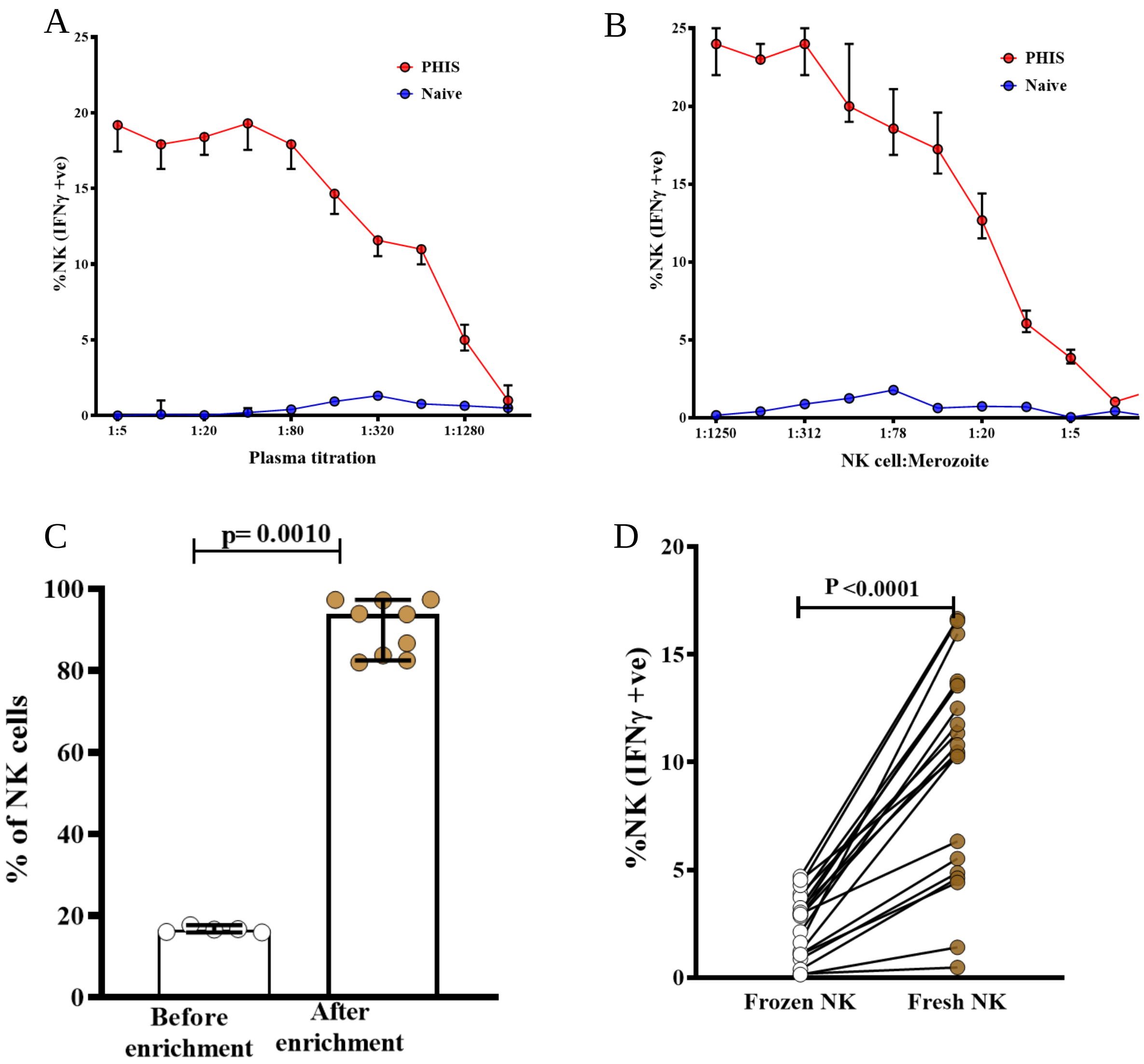


**Supplementary Figure 1. Optimization of novel ab-NK assay.** Dose-dependent, NK cell IFNγ production with increasing antibody concentration **(A)** and NK cell to merozoite ratio **(B)**. Merozoites were opsonized using a pool of hyperimmune serum (PHIS) from adults in Kilifi. Error bars represent 95% confidence interval (CI) of the median of three replicates. **(C)** The proportion of NK cells (CD56+, CD3-) before (n=5) and after (n=9) NK cell enrichment. Error bars represent 95% CI of the median; Mann-Whitney test. (**D**) Higher IFNγ production in NK cells isolated from fresh versus frozen PBMC. Plasma samples from Junju adults (N=25); P-value from the Wilcoxon matched-pairs signed rank test.

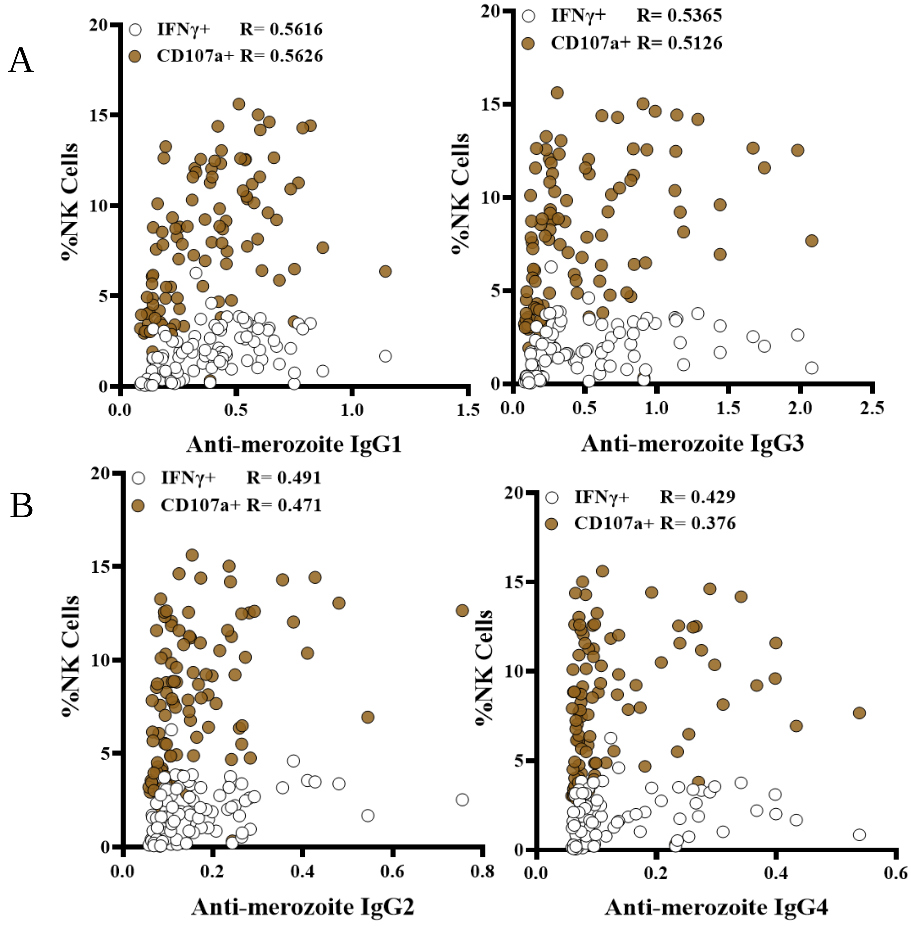


**Supplementary Figure 2. Ab-NK activity is correlated with anti-merozoite antibody binding** Two-way scatter plots demonstrate the correlation between Ab-NK cell degranulation and IFNγ production respectively, versus IgG binding antibodies against merozoites. Higher correlations observed for cytophilic (A & B) compared to non-cytophilic (C & D) isotypes. The x-axis shows the ELISA optical density; Spearman's R, $P < 0.001$ for all comparisons; n = 142 CHMI samples)

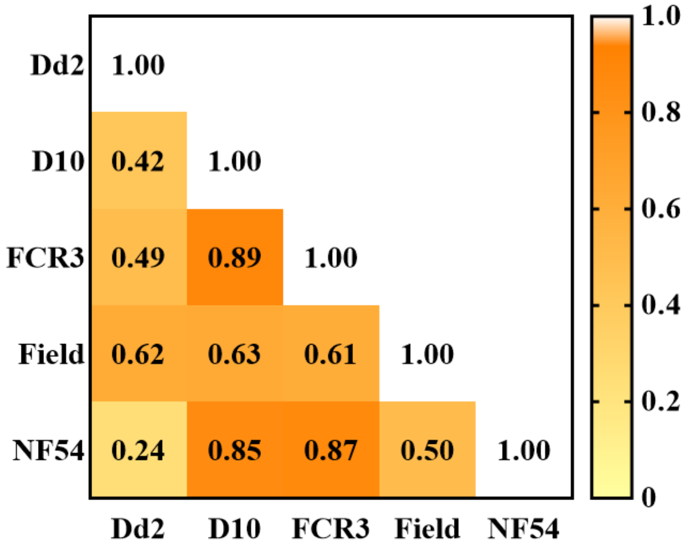


**Supplementary Figure 3. Ab-NK activity mediates strain transcending responses:** Spearman correlation heatmap between the proportion of NK cells producing IFNγ upon activation by opsonized merozoites from four *P. falciparum* strains of different geographical origin; West Africa (NF54) , Gambia (FCR3), Papuap New Gunea (D10) S.E Asia (Dd2) and a recently adapted clinical isolate from Kenya (Field; n=20).

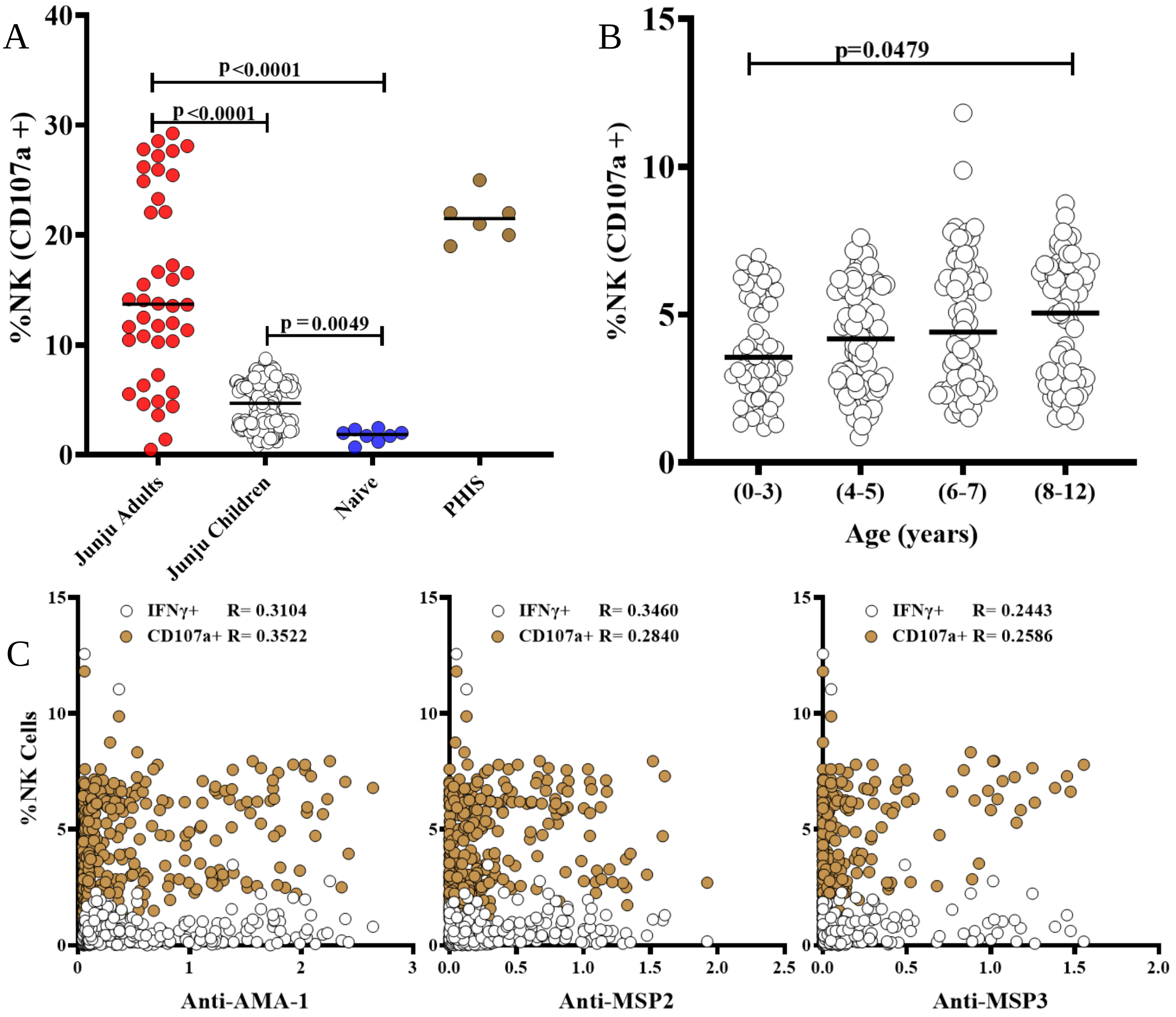


**Supplementary Figure 4. Antibody mediated NK cell activity in children:** (**A**) Merozoite opsonizing antibodies from Junju adults (n=40) induced significantly higher proportion of ab-NK cells degranulation compared to Junju children (n=293). Hyperimmune (PHIS) and malaria naive plasma were included as positive and negative controls respectively. Bars represents the median value; Kruskal-Wallis with Dunn's multiple comparison test. (**B**) Ab-NK cell degranulation in Junju children increased with age; Kruskal-Wallis with Dunn's multiple comparison test (n=293). (**C**) Spearman correlation between ab-NK cell degranulation or IFNγ production and total IgG ELISA responses against recombinant *P falciparum* AMA-1, MSP2 or MSP3 antigens (n=293).

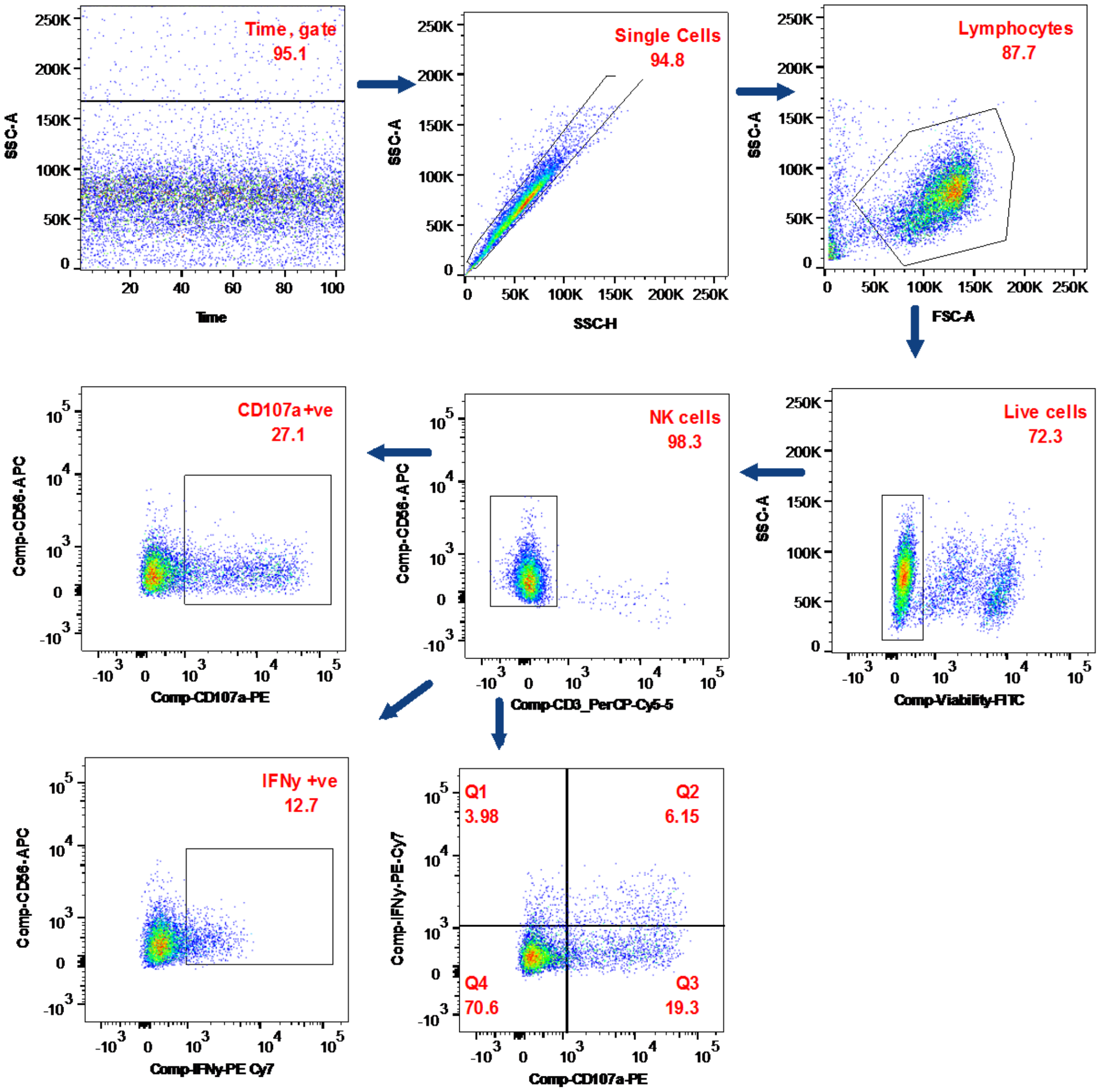


**Supplementary figure 5. Flow cytometry gating strategy**: The acquisition of stimulated NK cells data was shown using time and singlets excluding cell debri and double events. respectively. Lymphocytes were defined based on size and granularity before gating on live cells. Next, NK cell identification and function were identified, using the panel in

**Members of the CHMI-SIKA Study Team**

Abdirahman I Abdi[1], Yonas Abebe[3], Agnes Audi[4], Philip Bejon[1,2], Peter Billingsley[3], Peter C Bull[5], Primus Che Chi[1], Zaydah de Laurent[1], Susanne H Hodgson[6], Stephen Hoffman[3], Eric James[3], Irene Jao[1], Dorcas Kamuya[1], Gathoni Kamuyu[1], Silvia Kariuki[1], Nelson Kibinge[1],

Rinter Kimathi[1], Sam Kinyanjui[1,4,7], Cheryl Kivisi[7], Nelly Koskei[4], Mallika Imwong[8], Brett Lowe[1,2], Johnstone Makale[1], Kevin Marsh[1,2], Vicki Marsh[1,2], Khadija Said Mohammed[1], Moses Mosobo[1], Sean C Murphy[9], Linda Murungi[1], Jennifer Musyoki[1], Michelle Muthui[1], Jedidah Mwacharo[1], Daniel Mwanga[1], Joyce Mwongeli[1], Francis Ndungu[1], Maureen Njue[1], George Nyangweso[1], Domitila Kimani [1], Joyce M. Ngoi[1], Janet Musembi[1], Omar Ngoto[1], Edward Otieno[1], Bernhards Ogutu[4,11], Fredrick Olewe[4], James Oloo[4], Donwilliams Omuoyo[1], John Ongecha[4], Martin O Ongas[4,11], Michael Ooko[1], Jimmy Shangala[1], Betty Kim Lee Sim[2], Joel Tarning[2,12], Juliana Wambua[1], Thomas N Williams[1,13], Markus Winterberg[2,12]

Affiliations:

[1]KEMRI-Wellcome Trust Research Programme, Kilifi, Kenya

[2]Centre for Tropical Medicine and Global Health, Nuffield Department of Medicine, University Oxford, Oxford, UK

[3]Sanaria Inc., Rockville, MD, USA

[4]Centre for Clinical Research, Kenya Medical Research Institute, Kisumu, Kenya

[5]Department of Pathology, University of Cambridge, Cambridge, UK

[6]The Jenner Institute, University of Oxford, Oxford, UK.

[7]Pwani University, P. O. Box 195-80108, Kilifi, Kenya

[8]Faculty of Tropical Medicine, Department of Molecular Tropical Medicine and Genetics, Mahidol University, Bangkok, Thailand

[9]Departments of Laboratory Medicine and Microbiology, University of Washington, Seattle, Washington, USA

[10]Centre for Infectious Diseases, Heidelberg University Hospital, Heidelberg, Germany

[11]Center for Research in Therapeutic Sciences, Strathmore University, Nairobi, Kenya

[12]Mahidol-Oxford Tropical Medicine Research Unit, Mahidol University, Bangkok, Thailand

[13]Department of Medicine, Imperial College, London, UK
